# A model to estimate demand for personal protective equipment for Ontario acute care hospitals during the COVID-19 pandemic

**DOI:** 10.1101/2020.04.29.20085142

**Authors:** Kali Barrett, Yoshiko Nakamachi, Terra Ierasts, Yasin A. Khan, Stephen Mac, David Naimark, Nathan M. Stall, Raphael Ximenes, Andrew M. Morris, Beate Sander

## Abstract

In addition to instituting public health measures for COVID-19, managing healthcare resources is important for outcomes. The experiences in Italy and New York have shown that personal protective equipment (PPE) shortages can cause increased morbidity and mortality. We demonstrate a method to predict PPE demand across a health care system.

## Introduction

The novel coronavirus disease, COVID-19, caused by SARS-CoV2 has placed tremendous strain on populations and health care systems. Experiences in Italy and New York have demonstrated that a sufficient supply of personal protective equipment (PPE)–designed to protect health care workers and promote infection prevention and control—is one of the first vulnerabilities of the health care system when COVID-19 cases surge.[1] PPE shortages have added to the COVID-19 crisis globally, requiring health regions and hospitals to resort to extraordinary measures to source and procure supplies.[2] Predicting PPE needs is important to inform supply chain management and preserve adequate supplies. We used health system modelling to predict PPE demand in acute care settings, informed by interviews and direct observation of hospital administrators and healthcare workers caring for COVID-19 patients.

## Methods

We extend a previously described discrete time, dynamic, parallel, individual-level health state transition model that predicts COVID-19 emergency department (ED) visits and hospitalizations in Ontario (Canada’s largest province, population 14.6 million), based on observed pandemic trajectories in Ontario and other jurisdictions.[3] This model was designed to inform COVID-19 pandemic capacity planning in acute care. Based on the region’s population, the number of observed confirmed cases of COVID-19, and observed trajectories of case numbers, our model estimates the number of new cases of COVID-19 predicted to present to the ED for each day from March 6 to May 5, 2020. Cases are either sent home or admitted to hospital based on disease severity, and move through the acute care hospital system occupying ward or intensive care unit (ICU) beds, with or without invasive mechanical ventilation, based on probabilities derived from reported and observed data. We then estimated the number of suspected cases of COVID-19 (patients for whom PPE is required until COVID-19 infection is ruled out) by multiplying the projected number of confirmed cases by the ratio of confirmed to suspected cases currently reported to us by the Ontario Ministry of Health for patients in acute care (personal communication).

To determine the amount of PPE utilized per patient, we estimated patient “touchpoints”. A touchpoint was defined as any time a healthcare worker enters a patient room or is required to physically interact with a patient or their environment, during which PPE may be required. For each in-patient hospital setting (ED, ICU, and ward) we estimated the number of patient touchpoints within a 24-hour period, stratified by type of healthcare worker, COVID-19 status of the patient (confirmed, suspected or negative), and whether the patient receives invasive mechanical ventilation or is being turned prone.

We created “PPE bundles” for a confirmed case (COVID+), a person under investigation (PUI), or non-infected patients (COVID-) based on Public Health Ontario (PHO) guidance for PPE during the COVID-19 pandemic.[4] Table 1 lists the items of PPE included in each bundle. In brief, a PPE bundle for PUI or COVID+ patients requires a surgical mask, face shield, gown, and 1 pair of gloves. A PPE bundle for patients undergoing an aerosolizing generating medical procedure (AGMP) requires an N95 mask, extended length gloves and a face shield with neck drape. PPE consumption per patient was therefore calculated as the product of daily touchpoints and PPE bundles.

**Table 1.** Total PPE requirements for the Ontario acute care setting for suspected and confirmed COVID-19 patients

| Type of PPE | 10 day need around epidemic peak<br>(April 3-April 12) | 30 day need<br>(March 6-April 6) | 45 day need<br>(March 6-April 20) | 60 day need<br>(March 6-May 5) |
| --- | --- | --- | --- | --- |
| **Surgical mask** | 1,303,155 | 2,170,826 | 3,762,397 | 4,578,764 |
| **N95 mask** | 41,623 | 60,423 | 119,502 | 152,174 |
| **Gloves** | 3,688,383 | 5,815,652 | 10,726,711 | 13,572,199 |
| **Gloves (extended)** | 6,384 | 10,296 | 17,148 | 20,311 |
| **Face shield** | 1,341,587 | 2,226,102 | 3,873,325 | 4,720,782 |
| **Face shield with drape** | 3,192 | 5,148 | 8,574 | 10,156 |
| **Gown** | 1,344,779 | 2,231,250 | 3,881,899 | 4,730,938 |

Our model estimating predicted cases of COVID-19 was refined and recalibrated as additional empirical data became available so that case estimates were both consistent with past real-world cases while accurately predicting daily new cases. The estimated daily patient touchpoint counts were refined via interviews (with hospital service managers, point-of-care nurses, respiratory therapists, and physicians in critical care, general internal medicine, and the emergency department) and through direct observation at University Health Network and Sinai Health in Toronto, Canada. We identified that the use of N95 face masks was higher than expected based on PHO recommendations as a result of patient touchpoints for mechanically ventilated patients being cared for in the prone position. Patient care activities during patient proning, including turning to the prone position, turning back to the supine position, and the frequent head positioning changes, while not considered an AGMP by PHO, are considered potential AGMPs by local clinical leadership given the risk of ventilator circuit disconnection.[5] These care activities represented a significant source of N95 utilization. We adjusted our model to reflect this real-world practice. In the absence of high quality data to inform the probability of patients being proned, we assumed that 60% of mechanically ventilated patients were being proned based on the percentage of patients with a PaO2/FiO2 <150 mmHg reported in UK case series data.[6] We chose this PaO2/FiO2 threshold as it corresponds with both clinical guidelines for the care of patients with COVID-19 and clinical trial data.[5, 7]

Research ethics approval was obtained from the University of Toronto covering access to provincial databases, and was waived by the research ethics board at Sinai Health System for patient touchpoint data collection.

## RESULTS

We estimated that Ontario hospitals need 13.5 million gloves, 4.7 million face shields and gowns, 4.5 million surgical masks, and 152,174 N95 masks over a 60-day period near the height of the pandemic (Table 1). PPE demand varied according to the epidemic pattern and accompanying hospitalizations. It was highest around the peak of the pandemic and then declined in parallel with community prevalence.

## DISCUSSION

Our work shows that PPE demand can be predicted for a jurisdiction using a combination of epidemiologic projections, health system modelling, and hospital-specific data. It can be readily applied to other health care settings, and can be modified based on different policies within hospitals. Our methods of validation and calibration proved valuable, as initial estimates of PPE use were lower than expected, especially for N95 facemasks. Our work is strengthened by real-world observations of clinical volumes and practice patterns, and has directly informed PPE procurement activities by the Ministry of Health for the Province of Ontario.

A potential limitation of our study is we did not consider individual organizational PPE use policy such as universal masking and extended use or reuse of specific items of PPE such as face-shields. Our information-gathering activities to support this work revealed that PPE use guidelines varied between institutions, and while always based on healthcare worker safety, were influenced by factors including patient case volumes and local PPE stocks. We therefore chose to base our model on a non-restrictive model of PPE utilization. We argue that health systems should be planning their PPE needs based on non-restrictive, or “worst-case” estimates of PPE demand, in order to prevent both patient and health care worker morbidity and mortality.

It is possible that strategies such as universal masking, while often considered “conservation strategies” may in fact represent a significant source of PPE utilization as health systems resume previously suspended non-COVID-19 related clinical activities and there are greater numbers of healthcare workers working in acute health care systems in the weeks and months to come. We are developing an online tool on our website (www.covid-19-mc.ca) to help health systems estimate the amount of PPE required if conservation strategies are deployed. Specifically, organizations will be able to predict their PPE needs based on their local health human workforce data and conservation strategy type.

To our knowledge, this is the first study to estimate PPE need based on both dynamic modelling of projected cases of COVID-19, and episodes of patient contact. The US Centers for Disease Control and Prevention has released a PPE “burn rate calculator”, but it is a static model based on institutional PPE use and is neither informed by epidemiologic data nor patient and healthcare worker specific data.[8] We caution against estimating PPE demand based on so called “burn rates”, as many institutions have already depleted PPE stocks, and may be implementing conservation strategies such as reuse of masks or restricted distribution in an attempt to conserve supplies. Planning based on historical use may significantly underestimate true PPE needs—especially if case volumes rise—and may lead to inaccurate assumptions regarding future PPE needs that are insufficient to meet the needs of protecting healthcare workers.

Future work to better describe PPE utilization and explore the impact of restrictive measures of PPE use will allow for better estimates of PPE demands.

## CONCLUSION

Modelling based on epidemiologic data, and clinical practice patterns, is a data-driven method of estimating future PPE needs for a region. Our model demonstrates that PPE demand for the Province of Ontario is substantial and that current stockpiles may be insufficient to meet demand, potentially impacting the safety of healthcare workers, the sustainability of the workforce, and overall health outcomes for acute and critical care patients during the COVID-19 pandemic.

## Data Availability

Data regarding the health systems model is available at www.covid-19-mc.ca.

## REFERENCES

1. Ranney ML, Griffeth V, Jha AK. Critical Supply Shortages - The Need for Ventilators and Personal Protective Equipment during the Covid-19 Pandemic. N Engl J Med 2020.

2. Artenstein AW. In Pursuit of PPE. New England Journal of Medicine 2020: e46.

3. Barrett K, Khan YA, Mac S, Ximenes R, Naimark DMJ, Sander B. Potential magnitude of COVID-19-induced healthcare resource depletion in Ontario, Canada. medRxiv 2020.

4. Ontario Agency for Health Protection and Promotion (Public Health Ontario). IPAC recommendations for use of personal protective equipment for care of individuals with suspect or confirmed COVID-19. Toronto, Canada: Queens’s Printer for Ontario, 2020.

5. University of Toronto IDoCCM. Management Principals of Adult Critically Ill COVID-19 Patients. Available at: https://www.criticalcare.utoronto.ca/covid-19-resources. Accessed April 27.

6. Intensive Care National Audit & Research Centre. ICNARC report on COVID-19 in critical care, 2020 24 April.

7. Guerin C, Reignier J, Richard JC, et al. Prone positioning in severe acute respiratory distress syndrome. N Engl J Med 2013; 368(23): 2159–68.

8. Centers for Disease Control and Prevention. Personal Protective Equipment (PPE) Burn Rate Calculator. Available at: https://www.cdc.gov/coronavirus/2019-ncov/hcp/ppe-strategy/burn-calculator.html. Accessed April 27.

